# Tracheobronchial Stenosis in Granulomatosis with Polyangiitis: Immunosuppressant Use and Airway Dilation Frequency

**DOI:** 10.1101/2025.07.25.25332215

**Authors:** Brendan Denvir, Ami A. Shah, Alexander T. Hillel, Philip Seo, Ji Soo Kim, Brendan Antiochos

## Abstract

**Objective:** Tracheobronchial stenosis (TBS) occurs in 13–27% of patients with granulomatosis with polyangiitis (GPA) and may cause life-threatening airway compromise. Despite advances in treatment, TBS remains difficult to manage, with frequent relapses and high procedural burden. The objective of this study was to evaluate the relationship between immunosuppressant use and frequency of relapse in patients with TBS-GPA.

**Methods:** We performed retrospective review of patients with TBS-GPA seen at Johns Hopkins Medical Institutions between 2013-2024. Baseline demographic and clinical characteristics, immunosuppressant exposure, and tracheal dilation procedure dates were abstracted. A multivariate mixed effects Poisson regression model was used to assess the association between immunosuppressant exposures (rituximab, cyclophosphamide, methotrexate, azathioprine, leflunomide, and mycophenolate) and tracheal dilation incidence, adjusting for age, years since TBS diagnosis, anti-neutrophil cytoplasmic antibody (ANCA) status, GPA disease severity, and concomitant treatment with glucocorticoid injections.

**Results:** A total of 56 patients with TBS-GPA were included in the analysis, with a mean follow-up duration of 9.9 years. In the adjusted mixed-effects Poisson model, patient-years on leflunomide were associated with a 64% lower incidence of tracheal dilations compared to periods off leflunomide (IRR 0.36, p = 0.002). No statistically significant associations were observed for the other immunosuppressants measured. Among other tested covariates, age under 40, severe GPA, and concomitant glucocorticoid injections were associated with higher dilation frequency.

**Conclusion:** Leflunomide use was associated with a lower frequency of tracheal dilations in patients with TBS-GPA. These findings support the need for further evaluation of leflunomide as a treatment option in this population.

---

Granulomatosis with polyangiitis (GPA) is an anti-neutrophil cytoplasmic antibody (ANCA)-associated vasculitis characterized by widespread necrotizing granulomatous inflammation. While it can involve nearly all organ systems, GPA frequently involves the airway, affecting both upper and lower segments of the respiratory tract.

Tracheobronchial stenosis (TBS) is a potentially life-threatening manifestation of GPA in the respiratory tract that occurs in an estimated 13–27% of patients.^1–6^ Stenosis most commonly affects the subglottic region just below the larynx,^7^ though involvement of the larynx, lower trachea, and bronchi also occur.^8^

The management of tracheobronchial disease in GPA relies primarily on procedural intervention. Common approaches include balloon dilation, local corticosteroid injection, and surgical laryngotracheal resection and/or reconstruction.^9,10^ In severe cases, patients will require tracheostomy. Although these interventions serve to re-open the airway acutely, disease relapse after intervention is frequent. Observed relapse rates after procedural intervention vary by study between 38.5% and 83.3%, with the mean number of procedures per patient of 3.5.^9,11^ Tracheostomy is required in the disease course of 20–40% of patients with TBS-GPA.^1,12–14^

Unlike other GPA manifestations, the response of TBS to systemic immunosuppression remains poorly defined. Randomized controlled trials in GPA have not specifically evaluated the impact of treatment on tracheobronchial involvement, and existing observational studies have yielded mixed results. Some data suggest a potential benefit of systemic therapy: two observational studies reported lower relapse rates among patients who received immunosuppression compared to those who did not.^5,15^ A third retrospective cohort study found reduced relapse rates among patients treated with more than 30 milligrams of daily prednisone, however, they found no such effect for steroid-sparing immunosuppressants.^16^ Aden et al. observed lower recurrence rates in patients treated with rituximab compared to those treated with csDMARDs.^17^ In contrast, several cohorts have documented frequent isolated airway relapses in patients with otherwise well-controlled systemic disease, raising questions about the overall efficacy of systemic immunosuppression for TBS.^1,3^

In light of limited and conflicting evidence, the 2021 American College of Rheumatology/Vasculitis Foundation Guidelines recommend systemic immunosuppression for treatment of tracheobronchial disease in GPA, however, they do not specify a preferred agent.^18^

In this study, we sought to characterize the clinical features of patients with tracheobronchial GPA treated at our center and to evaluate the association between immunosuppressant use and the incidence of tracheobronchial disease relapse. This analysis builds on prior work from our institution,^19^ incorporating a larger patient cohort and extended follow-up time to provide a more comprehensive assessment of treatment outcomes. Based on prior studies and our center’s clinical experience, we hypothesized that leflunomide and rituximab would be associated with a lower incidence of tracheal dilations, reflecting a potential therapeutic advantage in treating this manifestation.

## Patients and Methods

### Patient Selection

We identified patients through two avenues (Figure 1). First, we queried Epic SlicerDicer for patients with ICD-10 codes associated with TBS (J38.6, J39.8, J98.09) who were seen by a Johns Hopkins rheumatologist between January 1, 2013 and December 31, 2024. This query was designed to capture patients with TBS who were seen by rheumatology, regardless of whether they had underlying GPA. This query yielded 106 patients. Our second query utilized the Johns Hopkins Precision Medicine Analytics Platform (PMAP) Vasculitis Registry. This is an IRB-approved registry database (IRB#00368120) that includes all patients with vasculitis related diagnoses that have been seen at Johns Hopkins Medical Institutions since January 1, 2013. We queried PMAP for patients with ICD-10 codes associated with GPA (M31.30, M31.31, I77.82) and TBS. This yielded 84 patients. After accounting for overlap between the two queries, we identified a total of 123 patients who were analyzed by retrospective chart review to confirm that each patient was diagnosed with GPA by a rheumatologist and that an otolaryngologist and/or pulmonologist made the diagnosis of TBS by laryngoscopy and/or bronchoscopy. Patients were excluded from the final analysis if: (i) they were not seen in Johns Hopkins Rheumatology, Otolaryngology, or Pulmonology Departments; (ii) there was no chart evidence of TBS attributed to GPA; or (iii) they had less than six months of follow-up data after their initial visit. Our final study population included 56 patients.

**Figure 1.**
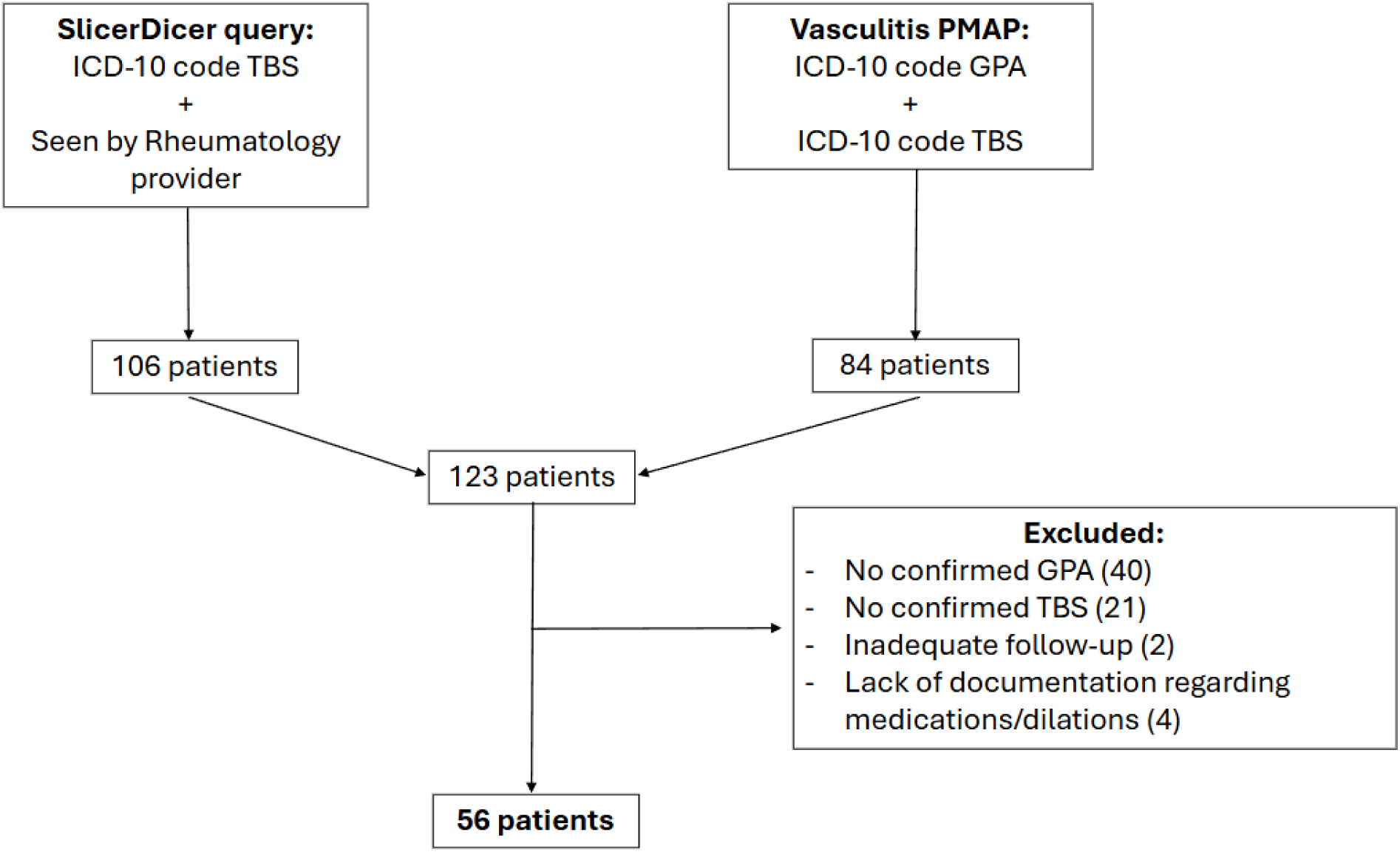
Patient Selection Flow Diagram. Separate queries were conducted using Epic SlicerDicer and the Vasculitis Precision Medicine Analytics Platform (PMAP). After excluding duplicate patients, a total of 123 unique patient charts were identified and reviewed.

### Data Collection

We used a standardized data extraction form in REDCap to record data for all patients who met inclusion and exclusion criteria. We collected baseline data on demographic characteristics, clinical features related to GPA, and characteristics of tracheobronchial stenosis. Demographic data included age, sex, and race. GPA-related clinic characteristics included the date of GPA diagnosis, serologies (ANCA, anti-MPO antibody, and anti-PR3 antibody), organ involvement at diagnosis, and biopsy reports of areas of disease involvement. Organ involvement was recorded using a list of organ-specific GPA disease manifestations adapted from the Birmingham Vasculitis Activity Score (BVAS) and Vasculitis Damage Index (VDI). TBS-related clinical characteristics included the date of TBS diagnosis, the presence of airway symptoms (dyspnea, stridor, dysphonia, hemoptysis), the anatomic location of airway stenoses, and airway biopsy results.

Immunosuppressant medication data were abstracted by reviewing rheumatology and otolaryngology notes. We documented start and stop dates associated with rituximab, cyclophosphamide, methotrexate, azathioprine, leflunomide, and mycophenolate mofetil. For rituximab specifically, the treatment “stop date” was determined to be 18 months after the last dose, reflecting its prolonged biologic effect in GPA.^20^ If a patient received consecutive doses within 18 months, then this was considered continuous treatment.

The dates and types of procedural interventions performed were recorded via review of otolaryngology, pulmonology, and procedural notes. Types of procedure included: tracheal or bronchial balloon dilation, laryngotracheoplasty, laryngotracheal resection surgery, and tracheostomy placement. When dilations occurred concurrently with local glucocorticoid injection and/or laser excision, these adjunctive procedures were documented. Mortality data, including dates and causes of death, were also documented.

The start of follow-up period for each patient was determined as either the time of tracheobronchial stenosis diagnosis, or the time after TBS diagnosis at which data regarding tracheal dilations and medications was available. Follow-up concluded at the patient’s last rheumatology or otolaryngology visit before December 31, 2024.

### Data Analysis

All analyses were conducted in R. Descriptive statistics were used to describe the demographics, GPA-related characteristics, and TBS-related characteristics of the cohort.

To enable longitudinal analysis, we transformed the dataset into a long format, with each row representing a distinct medication regimen interval. Whenever a medication change occurred, a new interval was initiated. Each interval included binary indicator variables denoting the use of specific immunosuppressants, the duration of the interval in days, and the total number of tracheal dilations performed during that interval. The primary outcome variable was the frequency of tracheal dilations. Additionally, each interval captured the elapsed time since the patient’s diagnosis of TBS.

We included several non-time-varying covariates in the analysis, specifically binary indicators for ANCA status (positive vs. negative), disease severity (severe vs. non - severe), age greater than 40 years at the time of TBS diagnosis, and concomitant treatment with intralesional glucocorticoid injections. ANCA positivity was defined by at least one positive result for p-ANCA, c-ANCA, anti-PR3, or anti-MPO antibodies at any point during follow-up. Severe disease classification required the presence of organ - threatening manifestations as defined by the ACR/VF guidelines including glomerulonephritis, mononeuritis multiplex, diffuse alveolar hemorrhage, gastrointestinal ischemia, or cardiac involvement.^18^

We used univariate Poisson regression models to evaluate the association between each predictor and the incidence of dilations. Subsequently, we developed a multivariate Poisson regression model that incorporated all covariates. We excluded medications used by fewer than five patients from this regression analysis. We included a random intercept term to account for patient-level variability in dilation frequency across follow-up time.

## Results

### Baseline Demographic and Clinical Characteristics

56 patients were included in the final analysis. The mean follow-up time was 9.9 years. Table 1 shows the baseline demographic and GPA related clinical characteristics, as well as an overview of immunosuppressant use of the cohort. The majority of patients were female (67.9%) and white (78.1%). The average age at TBS diagnosis was 36.1 years. Most (91.1%) patients had sinonasal involvement, while 53.6% had lung involvement and 17.9% had renal involvement. The majority of patients were ANCA positive (80.4%), with 60.7% being PR3-ANCA positive and 19.6% MPO-ANCA positive. During the follow-up period, rituximab was the most widely used immunosuppressant medication with 58.9% of patients receiving it. Regarding other immunosuppressants, 42.9% of patients received methotrexate, 28.6% received leflunomide, and 23.2% received azathioprine.

**Table 1.** Baseline demographic, clinical, and treatment characteristics of patients with tracheobronchial GPA.

|  | Overall<br>(n = 56) |
| --- | --- |
| **Sex** |  |
| Male, n (%) | 18 (32.1) |
| Female, n (%) | 38 (67.9) |
| **Age, mean (sd)** | 36.8 (16.4) |
| **Age > 40 at TBS, n (%)** | 29 (51.8) |
| **Serologies\*** |  |
| ANCA positive, n (%) | 45 (80.4) |
| PR3-ANCA, n (%) | 34 (60.7) |
| MPO-ANCA, n (%) | 11 (19.6) |
| **GPA Severity†** |  |
| Severe, n (%) | 12 (21.4) |
| Non severe, n (%) | 44 (78.6) |
| **Organ Involvement‡** |  |
| Sinonasal, n (%) | 51 (91.1) |
| Pulmonary, n (%) | 30 (53.6) |
| Renal, n (%) | 10 (17.9) |
| Neurologic, n (%) | 2 (3.6) |
| Ophthalmologic, n (%) | 6 (10.7) |
| Skin/Mucous membranes, n (%) | 17 (30.4) |
| General, n (%) | 24 (42.9) |
| **Medication¶** |  |
| Rituximab, n (%) | 33 (58.9) |
| Methotrexate, n (%) | 24 (42.9) |
| Leflunomide, n (%) | 16 (28.6) |
| Azathioprine, n (%) | 13 (23.2) |
| Cyclophosphamide, n (%) | 5 (8.9) |
| Mycophenolate mofetil, n (%) | 4 (7.1) |

\* PR3-ANCA positivity was defined as either a positive anti-PR3 antibody by ELISA or a positive c-ANCA by indirect immunofluorescence. MPO-ANCA positivity was defined as either a positive anti-MPO antibody or a positive p-ANCA.
† Severe disease defined by the presence of life or organ-threatening manifestations consistent with American College of Rheumatology/Vasculitis Foundation guidelines
‡ Organ involvement adapted from Birmingham Vasculitis Activity Score and Vasculitis Damage Index. General = arthralgia, arthritis, myalgia, fever (> 38C), or weight loss (> 2 kg).
¶ Medications received at any point during the follow-up period

Table 2 shows TBS related clinical characteristics of the cohort. Almost half (44.6%) of patients had TBS diagnosed at or before GPA diagnosis, while the remaining patients (55.4%) were diagnosed with TBS after GPA. Among the latter group of patients, the median time from GPA to TBS diagnosis was 4.7 years (range: 0.2 – 25.6). Of these patients, 71% were on immunosuppressant medications at the time of TBS diagnosis. Regarding airway symptoms, the most common was dyspnea, present in 76.8% of patients. A minority of patients (5.4%) were asymptomatic at presentation. Regarding location of airway involvement, 87.5% of patients had subglottic involvement, and 10.7% of patients had bronchial involvement. Airway biopsy reports were available for 21 patients in our cohort. 17 biopsies showed inflammation, while only two showed granulomatous inflammation, and zero showed vasculitis. Patients diagnosed with TBS after GPA were less likely to have inflammation on their biopsy report compared to those diagnosed with TBS before or at GPA diagnosis (57.1% vs. 92.9%, p<0.01).

**Table 2.** Clinical, anatomic, and histopathologic features of airway disease in tracheobronchial GPA cohort.

|  | Overall<br>n=56 | TBS at/before GPA<br>(n=25) | TBS after GPA<br>(n=31) | p-value* |
| --- | --- | --- | --- | --- |
| **Time from GPA to TBS,**<br>**years** (Median [Range]) |  | -0.2<br>[-10.9, 0] | 4.7<br>[0.2, 25.6] | n/a |
| **Airway Symptoms, n = 56** |  |  |  |  |
| Dyspnea, n (%) | 43 (76.8) | 18 (72.0) | 25 (76.8) | 0.66 |
| Cough, n (%) | 9 (16.1) | 4 (16.0) | 5 (16.1) | 1.00 |
| Stridor, n (%) | 24 (42.9) | 10 (40.0) | 14 (45.2) | 0.91 |
| Dysphonia, n (%) | 16 (28.6) | 8 (32.0) | 8 (25.8) | 0.83 |
| Hemoptysis, n (%) | 4 (7.1) | 2 (8.0) | 2 (6.5) | 1.00 |
| None, n (%) | 3 (5.4) | 1 (4.0) | 2 (6.5) | 1.00 |
| **Area of Involvement, n = 56** |  |  |  |  |
| Glottic, n (%) | 11 (19.6) | 5 (20.0) | 6 (19.4) | 1.00 |
| Subglottic, n (%) | 49 (87.5) | 21 (84.0) | 28 (90.3) | 0.69 |
| Lower Trachea, n (%) | 3 (5.4) | 2 (8.0) | 1 (3.2) | 0.58 |
| Bronchial, n (%) | 6 (10.7) | 4 (16.0) | 2 (6.5) | 0.39 |
| **Histopathology, n=21†** |  |  |  |  |
| Inflammation, n (%) | 17 (80.9) | 13 (92.9) | 4 (57.1) | <0.01 |
| Vasculitis, n (%) | 0 (0.0) | 0 (0.0) | 0 (0.0) | 0.19 |
| Granuloma, n (%) | 2 (9.5) | 2 (14.3) | 0 (0.0) | 0.45 |
| Necrosis, n (%) | 1 (4.8) | 1 (7.1) | 0 (0.0) | 0.45 |
| Fibrosis, n (%) | 7 (33.3) | 3 (21.4) | 4 (57.1) | 0.10 |
| Granulation Tissue, n (%) | 7 (33.3) | 5 (35.7) | 2 (28.6) | 0.74 |

\* Chi-square tests were used for group comparisons, with Fisher's exact test applied when expected cell counts were <5
† Biopsy findings were available in 21 patients: 14 in patients with TBS at or before GPA, and 7 in patients with TBS after GPA

### Mortality and Procedural Data

During the follow-up period, four patients died—three from COVID-19 and one from an unknown cause (Table 3). A total of 278 tracheal dilations were performed. Of these, 57.9% were performed with concomitant local steroid injections and 16.2% with concomitant excisions. The median number of dilations per patient was 3 (IQR 1–6.25), corresponding to a median rate of 0.36 dilations per year (IQR 0.26–0.85). Eleven patients did not undergo any dilation procedures. There were five tracheostomies during the follow-up period and an additional two patients had tracheostomies performed before the follow-up. Seven patients underwent laryngotracheal surgery during the follow-up period.

**Table 3.** Summary of Mortality and Procedural Data from tracheobronchial GPA cohort.

|  | Overall<br>(n=56) |
| --- | --- |
| **Number of Deaths** | 4 |
| COVID-19 | 3 |
| Unknown | 1 |
| **Total Dilations, n** | 278 |
| With intralesional steroid injection, n (%) | 161 (57.9) |
| With excision procedure, n (%) | 45 (16.2) |
| With mitomycin-C application, n (%) | 15 (5.4) |
| **Dilations per patient, median (IQR)** | 3.0 (1.0, 6.25) |
| **Dilations per patient per year, median (IQR)** | 0.36 (0.26, 0.85) |
| **Patients with Zero Dilations, n** | 11 |
| **Total Tracheostomies\*, n** | 5 |
| **Total Tracheal Surgeries†, n** | 7 |

\* Five tracheostomies occurred during the follow-up period. 2 additional patients had tracheostomies before the start of follow-up
† Tracheal surgeries include laryngotracheoplasty and tracheal resection

### Regression Analyses

Table 4 shows the results of univariate and multivariate regressions. In our multivariate model, among immunosuppressants, leflunomide was associated with reduced dilation incidence, while controlling for treatment with other medications as well as age, time since diagnosis, GPA disease severity, and ANCA status (IRR 0.36, p=0.002). No such association was observed for the other immunosuppressants included in the model (rituximab, methotrexate, and azathioprine). Among the other covariates, age over 40 at TBS diagnosis was associated with a lower dilation incidence (IRR 0.53, p=0.039). Patients with severe GPA had a higher incidence of tracheal dilation compared to non-severe patients (IRR 2.32, p=0.019), as well as patients receiving concomitant glucocorticoid injections (IRR 2.12, p<0.001).

**Table 4.** Incidence rate ratios estimated from univariate models and multivariate mixed effects Poisson regression model of tracheal dilation incidence.

|  | <u>Univariate Models</u> |  | <u>Multivariate model</u> |  |
| --- | --- | --- | --- | --- |
|  | IRR (95% CI) | p-value | IRR (95% CI) | p-value* |
| **Immunosuppressants** |  |  |  |  |
| Rituximab | 1.02 (0.69, 1.50) | 0.935 | 1.08 (0.72, 1.62) | 0.723 |
| Methotrexate | 1.68 (1.13, 2.49) | 0.010 | 1.33 (0.87, 1.99) | 0.178 |
| Leflunomide | 0.32 (0.18, 0.57) | <0.001 | 0.36 (0.19, 0.68) | 0.002 |
| Azathioprine | 0.91 (0.51, 1.62) | 0.752 | 0.80 (0.46, 1.39) | 0.423 |
| Cyclophosphamide † | 1.81 (0.28, 6.60) | 0.441 | - | - |
| Mycophenolate mofetil † | 0.67 (0.14, 2.49) | 0.580 | - | - |
| **Additional Covariates** |  |  |  |  |
| Years from TBS diagnosis | 0.95 (0.91, 0.98) | 0.003 | 0.97 (0.93, 1.00) | 0.052 |
| Age over 40 at TBS diagnosis | 0.64 (0.36, 1.16) | 0.135 | 0.53 (0.28, 0.97) | 0.039 |
| Severe disease ‡ | 2.16 (1.13, 4.23) | 0.019 | 2.32 (1.14, 4.85) | 0.019 |
| ANCA status ¶ | 3.10 (1.45, 6.89) | 0.004 | 2.22 (0.93, 5.45) | 0.071 |
| Concomitant steroid injections | 2.23 (1.50, 3.38) | <0.001 | 2.12 (1.40, 3.25) | <0.001 |

\*P-value less than 0.0125 is significant after Bonferroni correction adjusting for multiple comparisons
† Cyclophosphamide and mycophenolate mofetil were not included in the final multivariate model due to limited use in our sample (fewer than 6 patients on each medication), and poorer model performance with their inclusion.
‡ Severe GPA was defined by the presence of glomerulonephritis, diffuse alveolar hemorrhage, mononeuritis multiplex, gastrointestinal ischemia, or cardiac involvement. All other patients were non-severe.
¶ ANCA status was positive if c-ANCA, p-ANCA, anti-PR3 antibody, or anti-MPO antibody were positive at vasculitis diagnosis.

## Discussion

Tracheobronchial disease of GPA is a challenging and understudied manifestation, with limited evidence available to guide the use of systemic immunosuppression or the optimal choice of agent. In this context, we conducted a retrospective review of patients with TBS-GPA seen at our center over a 12-year period, evaluating the association between immunosuppressant use and the incidence of TBS relapse. Among the immunosuppressants analyzed, only leflunomide was associated with a reduced incidence of relapse. We also found that younger age, the presence of severe GPA, and concomitant glucocorticoid injections were associated with higher relapse rates. By leveraging a large cohort with extended follow-up, this study adds to the growing body of literature on clinical and treatment-related factors linked to disease activity in TBS-GPA. Notably, it includes the largest reported number of TBS-GPA patients treated with leflunomide to date. While these findings do not establish causality, they build on prior observations by Chen et al.^19^ and support further investigation into whether leflunomide may offer a therapeutic advantage in managing tracheobronchial GPA.

Over the past decade, mounting evidence from retrospective studies has suggested that systemic immunosuppression may reduce relapse rates in patients with TBS-GPA. In 2015, Girard et al. reported a 100% relapse rate among patients treated with local therapy alone, compared to 59% among those who received both local and systemic therapy.^15^ Similarly, Moroni et al. (2023) retrospectively evaluated 21 patients with GPA and subglottic stenosis, finding a 100% relapse rate in the five patients who did not receive immunosuppression, compared to 44% in the 16 who did.^5^

Other studies have sought to compare the effectiveness of specific agents. Terrier et al. (2015) examined the relapse risk associated with several treatments and found no significant effect of csDMARDs, cyclophosphamide, or rituximab, though high -dose prednisone was associated with reduced relapse.^16^ Notably, only 6% of patients in that cohort received rituximab, limiting the power to detect a treatment effect. More recently, Aden et al. retrospectively compared relapse rates among patients with GPA and subglottic stenosis undergoing CO₂ laser excision. Among 26 patients, they found lower relapse rates in those treated with concomitant rituximab (n=16) compared to those treated with csDMARDs (n=10).^17^ This was the first study, to our knowledge, to suggest superior efficacy of rituximab in TBS-GPA.

Similar to rituximab, the associations observed with glucocorticoid injections and methotrexate likely reflect confounding by indication. Glucocorticoid injections were associated with a higher incidence of tracheal dilations, which may indicate that they were more commonly administered to patients with more severe or refractory disease. Likewise, methotrexate use was associated with increased dilation frequency in univariate models; however, this association did not persist in multivariable analysis. Compared to leflunomide, methotrexate may have been preferentially used in patients with more severe disease, potentially explaining the observed differences. These findings highlight the challenge of disentangling treatment effect from disease severity in observational studies of TBS-GPA.

Although no studies to date have directly compared the efficacy of different csDMARDs specifically for TBS-GPA, several studies have evaluated csDMARDs in the broader context of GPA maintenance therapy. Leflunomide has been proposed as a potentially superior option based on a 2014 network meta-analysis that indirectly compared leflunomide, methotrexate, azathioprine, and mycophenolate mofetil.^21^ This analysis synthesized results from three trials: methotrexate vs. leflunomide, methotrexate vs. azathioprine, and azathioprine vs. mycophenolate mofetil. Among these comparisons, leflunomide was associated with the highest relapse-free survival, achieving statistical significance at the 90% confidence level.^21^

This paper, in conjunction with the findings in our study, led us to consider whether the mechanistic properties of leflunomide might account for its observed clinical benefit in this setting. It has been suggested that treatment resistance in tracheobronchial disease may be driven by irreversible fibrotic remodeling rather than persistent active inflammation.^6,16,22–25^ Notably, leflunomide has demonstrated anti-fibrotic effects in several animal models, including inhibition of fibrogenic cytokines and intracellular signaling pathways, and induction of apoptosis in fibrogenic cells.^26–28^ These mechanistic features, taken together with our clinical findings, support the hypothesis that leflunomide may have a unique therapeutic role in targeting fibrotic airway disease in GPA.

Similar to rituximab, we suspect that confounding by indication is at play in the findings associated with concomitant glucocorticoid injections and methotrexate. While we found that glucocorticoid injections were associated with higher incidence of dilation procedures, this may reflect that patients with more refractory disease were treated more aggressively with concomitant injections. Similarly, in univariate modeling, methotrexate was associated with higher dilation incidence. Compared to leflunomide, methotrexate may be more commonly used in severe patients which accounts for this finding, and accounts for resolution of the effect in multivariate modeling.

The demographic characteristics of our cohort are broadly consistent with those reported in prior observational studies. Our population was predominantly female and white, reflecting the known demographic distribution of GPA.^2,3,13,16^ We observed a wide range of ages at tracheobronchial disease diagnosis, consistent with prior reports.^1,3,16^ Additionally, the timing of TBS diagnosis relative to GPA diagnosis varied substantially, a pattern also noted in prior cohorts.^15^ We found that younger age at diagnosis of TBS was associated with a higher incidence of relapse. To our knowledge, this has not been reported in previous literature regarding tracheobronchial disease specifically, but is corroborated by prior research linking younger age at diagnosis with more severe or relapsing forms of ANCA-associated vasculitis.^30^ Additionally, Bohm et al. reported that SGS was more common among pediatric-onset GPA in their cohort compared to adult-onset GPA.^31^

GPA-related clinical features in our cohort were also consistent with existing literature. A minority of patients had severe organ involvement, including renal disease. Prior studies report variable rates of renal involvement in TBS-GPA, ranging from 28% to 53%.^1,3,16^ Reported rates of ANCA-negative disease range from 6% to 25% across studies.^1–4,16^ We suspect our cohort includes a relatively high proportion of patients with non-severe and ANCA-negative disease due to our center’s close collaboration with ENT providers and, therefore, high proportion of referrals of isolated otolaryngologic disease.

With respect to baseline characteristics of TBS, our cohort was similar to previously reported cohorts in terms of airway involvement and biopsy findings. Subglottic involvement was present in the vast majority of patients, consistent with prior studies.^2,3,15,16^ Tissue biopsies primarily demonstrated non-specific inflammation, aligning with other reports in which vasculitis and/or granulomas are identified in only a minority of cases.^2,13,17^ We suspect that inflammation was less commonly reported on biopsies from patients with TBS diagnosed later in their disease course due to longer exposure to immunosuppression.

Finally, we found that the presence of extra-tracheobronchial organ-threatening disease was associated with a higher incidence of tracheal dilations. While both our data and prior studies indicate that TBS frequently occurs in patients with non -severe or isolated otolaryngologic GPA, this finding suggests an additional possibility: that patients with more severe systemic disease may also experience more severe, treatment-refractory TBS.

Several limitations should be considered when interpreting the findings of this study. First, this is a retrospective observational analysis conducted at a tertiary academic referral center, which may introduce selection bias toward patients with more severe or treatment-refractory disease. As such, the generalizability of our findings to individuals with milder tracheobronchial involvement or those managed in community settings may be limited. Second, information bias is an inherent limitation of studies relying on retrospective abstraction of EHR data. Variability in clinical documentation across providers and incomplete records may have led to misclassification of exposures, outcomes, or covariates. We did not effectively capture prednisone use in our cohort or stand-alone in-office intralesional steroid injections, both of which may have served as unmeasured confounding variables in our analysis. Third, our relatively small sample size limited statistical power and constrained our ability to adjust for confounding or apply more sophisticated causal inference techniques, such as propensity score matching, to estimate treatment effects more accurately. Future studies with larger, multicenter cohorts and prospective data collection will be essential to validate our findings and provide a more robust assessment of the comparative effectiveness of immunosuppressive therapies in tracheobronchial GPA.

Despite these limitations, this study has several notable strengths. First, it represents one of the largest cohorts of patients with tracheobronchial GPA reported to date, a noteworthy accomplishment given the rarity of this disease. Patients were followed for an average of 9.9 years, providing a longitudinal window sufficient to assess changes in dilation frequency over time. Such extended follow-up is essential for capturing the natural history of tracheobronchial involvement, which often progresses slowly and unpredictably. This prolonged observation period enhances our ability to detect the long-term effects of immunosuppressive therapies—effects that may take months to years to become apparent. Second, despite the modest sample size, we were able to adjust for key covariates, including GPA disease severity, ANCA status, time since TBS, and age in our regression models. Additionally, by incorporating random effects, we accounted for between-patient variability in procedural burden. These measures helped to mitigate important sources of confounding in our analysis.

This study advances our understanding of risk factors for severe, relapsing tracheobronchial disease in GPA and highlights a potential role for immunosuppressant choice—particularly leflunomide—in influencing disease trajectory. Future research should evaluate differences in disease course by age, as well as clinical and pathologic features distinguishing treatment responders from non-responders. Multicenter observational studies with larger, more diverse cohorts and methods to address treatment selection bias, such as propensity score matching, will be critical. In parallel, prospective studies are needed to assess the impact of specific therapies on objective pulmonary outcomes, including peak expiratory flow and airway imaging. Together, these efforts may inform more effective, individualized approaches to managing tracheobronchial GPA.

## Data Availability

All data produced in the present study are available upon reasonable request to the authors

## References

1. Langford, C. A. et al. Clinical features and therapeutic management of subglottic stenosis in patients with Wegener’s granulomatosis. Arthritis Rheum. 39, 1754–1760 (1996).

2. Marroquín-Fabián, E., Ruiz, N., Mena-Zúñiga, J. & Flores-Suárez, L. F. Frequency, treatment, evolution, and factors associated with the presence of tracheobronchial stenoses in granulomatosis with polyangiitis. Retrospective analysis of a case series from a single respiratory referral center. Semin. Arthritis Rheum. 48, 714–719 (2019).

3. Quinn, K. A. et al. Subglottic stenosis and endobronchial disease in granulomatosis with polyangiitis. Rheumatology 58, 2203–2211 (2019).

4. Martinez Del Pero, M., Jayne, D., Chaudhry, A., Sivasothy, P. & Jani, P. Long-term outcome of airway stenosis in granulomatosis with polyangiitis (Wegener granulomatosis): an observational study. JAMA Otolaryngol.--Head Neck Surg. 140, 1038–1044 (2014).

5. Moroni, L. et al. Role of systemic immunosuppression on subglottic stenosis in granulomatosis with polyangiitis: Analysis of a single-centre cohort. Eur. J. Intern. Med. 114, 108–112 (2023).

6. Hoffman, G. S. et al. Wegener granulomatosis: an analysis of 158 patients. Ann. Intern. Med. 116, 488–498 (1992).

7. Seth, R. & Alam, D. S. in.

8. Polychronopoulos, V. S., Prakash, U. B. S., Golbin, J. M., Edell, E. S. & Specks, U. Airway involvement in Wegener’s granulomatosis. Rheum. Dis. Clin. North Am. 33, 755–775, vi (2007).

9. Krol, R. M. et al. Systemic and Local Medical or Surgical Therapies for Ear, Nose and/or Throat Manifestations in ANCA-Associated Vasculitis: A Systematic Literature Review. J. Clin. Med. 12, 3173 (2023).

10. Carnevale, C., Arancibia-Tagle, D., Sarría-Echegaray, P., Til-Pérez, G. & Tomás-Barberán, M. Head and Neck Manifestations of Granulomatosis with Polyangiitis: A Retrospective analysis of 19 Patients and Review of the Literature. Int. Arch. Otorhinolaryngol. 23, 165–171 (2019).

11. Catano, J. et al. Presentation, Diagnosis, and Management of Subglottic and Tracheal Stenosis During Systemic Inflammatory Diseases. Chest 161, 257–265 (2022).

12. Taylor, S. C., Clayburgh, D. R., Rosenbaum, J. T. & Schindler, J. S. Clinical manifestations and treatment of idiopathic and Wegener granulomatosis-associated subglottic stenosis. JAMA Otolaryngol.--Head Neck Surg. 139, 76–81 (2013).

13. Gluth, M. B., Shinners, P. A. & Kasperbauer, J. L. Subglottic stenosis associated with Wegener’s granulomatosis. The Laryngoscope 113, 1304–1307 (2003).

14. Solans-Laqué, R. et al. Clinical features and therapeutic management of subglottic stenosis in patients with Wegener’s granulomatosis. Lupus 17, 832–836 (2008).

15. Girard, C., et al. Tracheobronchial Stenoses in Granulomatosis With Polyangiitis (Wegener’s): A Report on 26 Cases. Medicine (Baltimore) 94, e1088 (2015).

16. Terrier, B. et al. Granulomatosis with polyangiitis: endoscopic management of tracheobronchial stenosis: results from a multicentre experience. Rheumatol. Oxf. Engl. 54, 1852–1857 (2015).

17. Aden, A. A. et al. Medical Maintenance Therapy Following Laser Excision in Patients With Granulomatosis With Polyangiitis (GPA)-Associated Subglottic Stenosis. Otolaryngol.--Head Neck Surg. Off. J. Am. Acad. Otolaryngol.-Head Neck Surg. 171, 180–187 (2024).

18. Chung, S. A. et al. 2021 American College of Rheumatology/Vasculitis Foundation Guideline for the Management of Antineutrophil Cytoplasmic Antibody-Associated Vasculitis. Arthritis Care Res. 73, 1088–1105 (2021).

19. Chen, L. W. et al. Factors Affecting Dilation Interval in Patients With Granulomatosis With Polyangiitis-Associated Subglottic and Glottic Stenosis. Otolaryngol.--Head Neck Surg. Off. J. Am. Acad. Otolaryngol.-Head Neck Surg. 165, 845–853 (2021).

20. Thiel, J. et al. B cell repopulation kinetics after rituximab treatment in ANCA-associated vasculitides compared to rheumatoid arthritis, and connective tissue diseases: a longitudinal observational study on 120 patients. Arthritis Res. Ther. 19, 101 (2017).

21. Hazlewood, G. S. et al. Non-biologic remission maintenance therapy in adult patients with ANCA-associated vasculitis: a systematic review and network meta-analysis. Joint Bone Spine 81, 337–341 (2014).

22. Ugan, Y., Doğru, A., Aynalı, G., Şahin, M. & Tunç, Ş. E. A clinical threat in patients with granulomatosis polyangiitis in remission: Subglottic stenosis. *Eur*. J. Rheumatol. 5, 69–71 (2018).

23. Ye, W. et al. Characterizing the Cellular Constituents of Proximal Airway Disease in Granulomatosis With Polyangiitis. Otolaryngol.--Head Neck Surg. Off. J. Am. Acad. Otolaryngol.-Head Neck Surg. 172, 2009–2017 (2025).

24. Stoelben, E. [Idiopathic Subglottic Tracheal Stenosis]. Zentralbl. Chir. 149, 308–314 (2024).

25. Daum, T. E. et al. Tracheobronchial involvement in Wegener’s granulomatosis. Am. J. Respir. Crit. Care Med. 151, 522–526 (1995).

26. Yao, H., Li, J., Chen, J. & Xu, S. Inhibitory effect of leflunomide on hepatic fibrosis induced by CCl4 in rats. Acta Pharmacol. Sin. 25, 915–920 (2004).

27. Tang, X., Yang, J. & Li, J. Sensitization of human hepatic stellate cells to tumor necrosis factor-related apoptosis-inducing ligand-induced apoptosis by leflunomide. Biol. Pharm. Bull. 32, 963–967 (2009).

28. Morin, F. et al. Leflunomide prevents ROS-induced systemic fibrosis in mice. Free Radic. Biol. Med. 108, 192–203 (2017).

29. Terrier, B. et al. Granulomatosis with polyangiitis according to geographic origin and ethnicity: clinical-biological presentation and outcome in a French population. Rheumatol. Oxf. Engl. 56, 445–450 (2017).

30. Bloom, J. L. et al. The Association Between Age at Diagnosis and Disease Characteristics and Damage in Patients With ANCA-Associated Vasculitis. Arthritis Rheumatol. Hoboken NJ 75, 2216–2227 (2023).

31. Bohm, M. et al. Clinical features of childhood granulomatosis with polyangiitis (wegener’s granulomatosis). Pediatr. Rheumatol. Online J. 12, 18 (2014).

